# Hawkes process modeling of COVID-19 with mobility leading indicators and spatial covariates

**DOI:** 10.1101/2020.06.06.20124149

**Authors:** Wen-Hao Chiang, Xueying Liu, George Mohler

## Abstract

Hawkes processes are used in machine learning for event clustering and causal inference, while they also can be viewed as stochastic versions of popular compartmental models used in epidemiology. Here we show how to develop accurate models of COVID-19 transmission using Hawkes processes with spatial-temporal covariates. We model the conditional intensity of new COVID-19 cases and deaths in the U.S. at the county level, estimating the dynamic reproduction number of the virus within an EM algorithm through a regression on Google mobility indices and demographic covariates in the maximization step. We validate the approach on both short-term and long-term forecasting tasks, showing that the Hawkes process outperforms several benchmark models currently used to track the pandemic, including an ensemble approach and an SEIR-variant. We also investigate which covariates and mobility indices are most important for building forecasts of COVID-19 in the U.S.

## 1. Introduction

Mathematical modeling and forecasting are playing a pivotal role in the ongoing SARS-CoV-2 (COVID-19) pandemic. In mid-March 2020, a report out of Imperial College London [1] forecasted severe consequences in the U.S. and U.K. without significant public health interventions. In both nations, governments responded by closing schools, non-essential businesses and releasing general stay-at-home (shelter-in-place) orders. In the U.S., state and local policymakers are using mathematical models and projections to inform decisions about when and how to relax public health measures that have been put in place. By and large, compartmental models that explicitly incorporate transmission characteristics of infectious diseases have been favored over machine learning approaches. High profile Susceptible-Exposed-Infected-Removed (SEIR) models include those out of Columbia University [2], MIT [3], The Johns Hopkins University [4], and UCLA [5] (in the case of the UCLA model, an SEIR-variant with an unreported compartment is fit using least-squares to reported infection and recovery data). A major exception is the well-known IHME model [6], which employs Gaussian curve fitting to COVID-19 case and death count time series in locations further along (e.g., China, Europe) to estimate curves in locations where the outbreak is more recent (e.g., the United States). The IHME model has been called into question by epidemiologists because it lacks explicit transmission dynamics in the model [7]. Our goal in this paper is to show that Hawkes processes, widely used in the machine learning community to model contagion patterns in event data, are well suited for modeling and forecasting COVID-19 case and mortality data. They have several advantages that we will highlight, including being highly flexible in accommodating auxiliary spatio-temporal features such as county-level demographics and temporal mobility patterns, yet mathematically they are connected to compartmental models [8] and allow for explicit incorporation of transmission dynamics (which we briefly review in the following section). Furthermore, extensive research has been conducted in the past several years on a couple of machine learning techniques with the point process framework. Non-parametric Hawkes processes can be constructed where the triggering kernel is learned [9] and more recently, fully neural network based point processes have been developed [10, 11]. Sparse linear combinations of Hawkes processes were a winning solution in the 2017 NIJ Crime Forecasting Challenge [12]. In certain cases a mixture of Hawkes processes may be needed to model more complex event contagion with high dimensional marks through Dirichlet processes [13, 14]. Hawkes processes can also be used for causal inference on networks [15] and recent efforts have also focused on training point processes through reinforcement learning [16, 17]. We believe all of these methods have potential applications to modeling infectious diseases that are highly complex due to heterogeneity in the population, environment, and disparate public policies across regional and local jurisdictions. Despite these advantages, to our knowledge, the only U.S. state where a Hawkes process is being used to inform COVID-19 policy is in New Jersey (a collaboration with Facebook AI Research) ^1^.

The outline of the paper is as follows. In Section 2, we introduce our Hawkes process model whose productivity (reproduction number) is dynamic and depends on spatio-temporal covariates. Unlike recently introduced models that incorporate covariates into the background rate of a Hawkes process [18, 19], our Hawkes process model may be viewed as a convolution of lagged mobility with an inter-infection time distribution to estimate the intensity of secondary infections in the future. This is important as phased reopening in the U.S. leads to mobility changes, the effects of which are not realized in the case and mortality data until days or weeks later. Hence the model can be used to forecast changes in transmission and new cases in real-time as mobility changes (see Figure 1). We estimate the intensity along with the dynamic reproduction number of the virus within an EM algorithm through a regression on Google mobility indices and demographic covariates in the maximization step. In Section 3, we validate the approach on both short-term and long-term forecasting tasks, showing that the Hawkes process outperforms several benchmark models from “The COVID-19 Forecast Hub Network ^2^.” The benchmark models include SEIR models from the Columbia University [2], Johns Hopkins University Applied Physics Lab [4], and an ensemble model from Berkeley that uses combined linear and exponential predictors with spatial covariates [20]. We also investigate which covariates and mobility indices are most important for building forecasts of COVID-19 in the U.S. In Section 4, we discuss directions for future research and how the machine learning community may be able to help improve Hawkes process models of COVID-19 as the pandemic continues to unfold.

**Figure 1:**
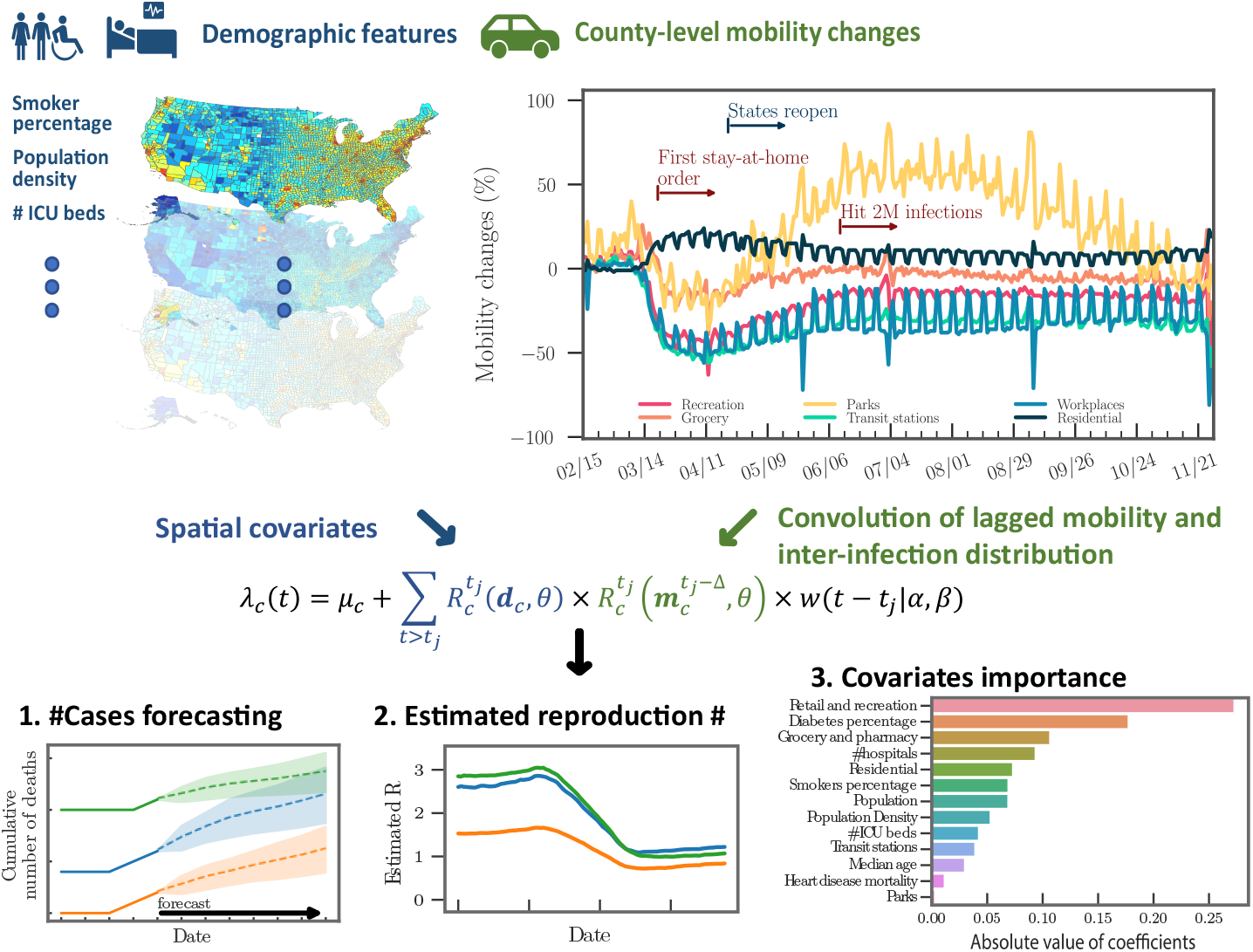
The framework of the Hawkes process model for COVID-19 transmission. Demo-graphic features at the county level impact the reproduction number of the Hawkes process. Lagged changes in mobility impact future secondary infections through a convolution with the inter-infection distribution *w*(*t*). The output of the model includes: (1) forecasts of future cases and mortality through simulation of the Hawkes process intensity, (2) an estimate of the dynamic reproduction number of the virus, and (3) regression results that allow for interpretation of the covariates that influence transmission differences across counties.

## 2. Hawkes process model of COVID-19 transmission

In this section, we introduce a Hawkes process with spatio-temporal covariates for modeling COVID-19 case and death data. We then discuss the connection of the model to compartment models used in epidemiology and develop an expectation-maximization algorithm for inference.

### 2.1. Incorporating covariates into the Hawkes process

We propose a novel Hawkes process model that simultaneously estimates the intensity of events and tracks the dynamic reproduction number of the virus. Given the timestamps (or dates), *𝒯* = {*t*_1_, *t*_2_, … *t*_*n*_}, of daily reported positive test cases or deaths, we model the rate of new cases (or deaths) in each country *c* as follows:

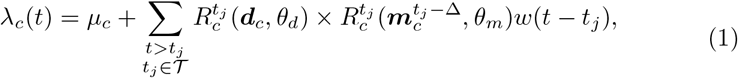

where *µ*_*c*_ is the background rate modeling imported infections, *w*(*t*) is the inter-infection time distribution, 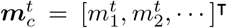 are mobility indices on day *t*, and ***d***_*c*_ = [*d*_1_, *d*_2_, *…*]^T^ are static demographic features. The time-varying reproduction number 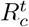 can be interpreted as the average number of secondary infections caused by a primary infection. Because we are modeling reported infections rather than time of exposure, we introduce the parameter Δ to capture a potential lag between a mobility change and the time *t*_*j*_ of a reported primary infection.

We then model the dynamic reproduction number 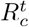through a Poissonregression,

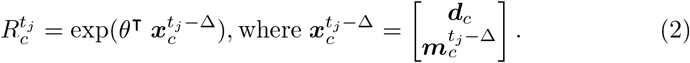

Here we have combined the spatial and temporal covariates to simplify notation in the rest of the paper. Our approach is related to those in recent preprints that incorporate mobility into compartment models [21, 4], however those approaches typically involve large-scale Monte Carlo simulations when performing inference. As we will show, the Hawkes process likelihood can be maximized without simulation via an efficient expectation-maximization algorithm.

### 2.2. Mathematical connection between Hawkes processes and compartmental models

Here we briefly review several variations of the Hawkes process in Equation 1 that can be connected to SEIR-type compartment models. The first variant is the SIR-Hawkes process. This model captures the long-term evolution of a pandemic by incorporating a pre-factor that accounts for the dynamic decrease in the number of susceptible individuals [8]:

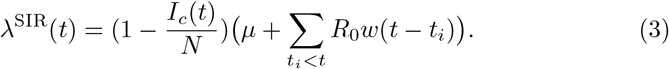

Here *I*_*c*_(*t*) is the cumulative number of infections that have occurred up to time *t* and *N* is the total population size. The point process governed by Equation 3 is a continuous time analog of a discrete stochastic SIR model when *w*(*t*) is specified to be exponential [8]. When *w*(*t*) is chosen to be gamma distributed, the Hawkes process also can approximate staged compartment models, like SEIR, if the average waiting time in each compartment is equal [22]. More complex parametric (or non-parametric) inter-infection time distributions *w*(*t*) may be employed within the Hawkes process framework in situations where disease dynamics cannot be captured by a SIR or SEIR model. In the early exponential growth stage of an epidemic, before finite population effects play a role (which is the case with current U.S. data), the Hawkes process in Equation 1 without the prefactor can be used to model new infections arising from SIR and SEIR models, as 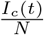 will be small.

While a pre-factor in the Hawkes process involving the cumulative number of infections, i.e., *I*_*c*_(*t*) in Equation 3, it is necessary to model long-term disease dynamics [8]. In the early stages of transmission, a linear Hawkes process can be used (as the prefactor will be close to 1),

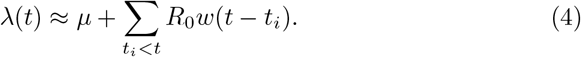

To illustrate this, we simulate a SEIR differential equation as the following:

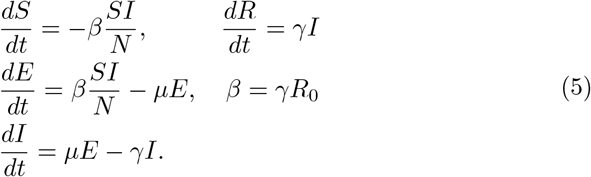

The parameters are chosen similar to those of COVID-19: *γ* = .1, *R*_0_ = 2, *µ* = 1, and *N* = 5·10^8^. We then fit the linear Hawkes process model in Equation 4 to new infections, *µE*, generated by the SEIR model. We use a non-parametric histogram estimator for *w*(*t*) and find a close fit between the Hawkes process and the SEIR model in Figure 2.

**Figure 2:**
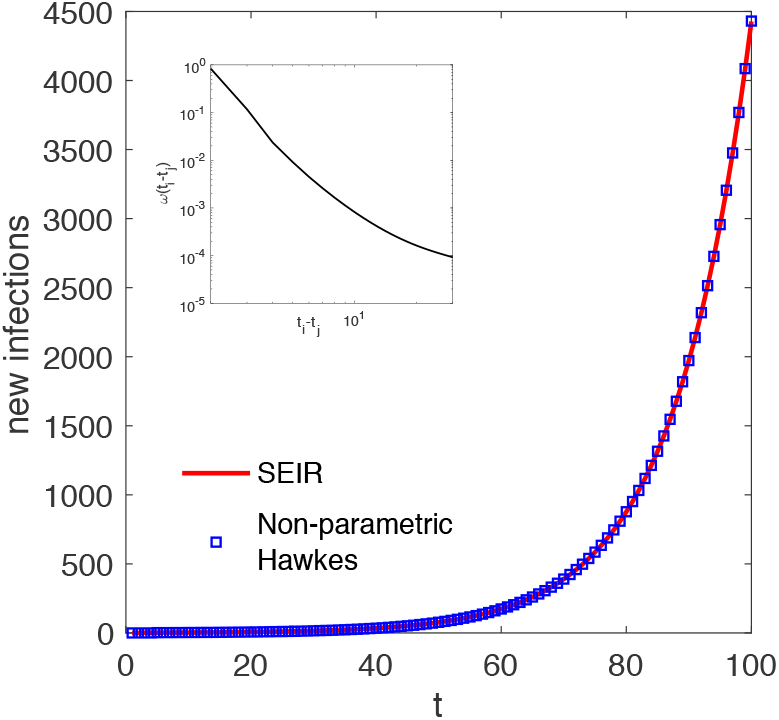
(Main figure) The red plot shows SEIR differential equation 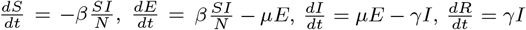, where *β* = *γR*_0_, *γ* = .1, *R*_0_ = 2, *µ* = 1, and *N* = 5 · 10^8^. The blue squares show that linear Hawkes process 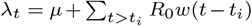 fit to the SEIR curve of new infections. Inset: Non-parametric histogram estimate for *w*(*t*).

### 2.3. EM algorithm for parameter inference

We use an expectation–maximization (EM) algorithm to estimate the model in Equation 1. First, we introduce latent random variables, *p*_*c*_(*i, j*), that represent the event that secondary infection *i* is caused by primary infection *j* in county *c*. We let *p*_*c*_(*i, i*) represent the event that case *i* is imported. The complete data log-likelihood is then given by,

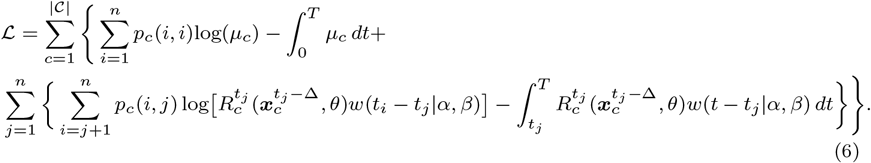

Here we use a Weibull distribution with shape *α* and scale *β* to model inter-infection times, which is used in other studies of epidemics [23, 24, 25] and we find accurately captures transmission in the present data [26].

As the branching structure of the process is unobservable, we optimize the complete data log-likelihood in Equation 6 by iteratively alternating between an expectation step where the branching probabilities *p*_*c*_ are estimated and a maximization step where model parameters are updated by maximizing Equation 6. The EM-algorithm is equivalent to a projected gradient ascent on the likelihood of the Hawkes process [27].

#### 2.3.1. Expectation step

During the expectation step, we estimate the latent variables *p*_*c*_(*i, j*) for each county. Given the parameters *θ, α, β*, and *µ*_*c*_ estimated from the last iteration, the probabilities that case *i* was caused by case *j* (Equation 7a) or was imported (Equation 7b) are given by:

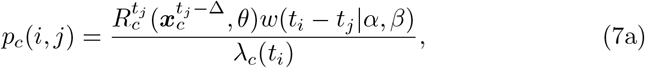

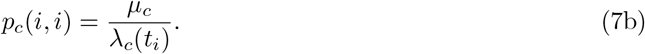

Note that the rate *λ*_*c*_(*t*_*i*_) in Equation 1 is considered to be an aggregation of triggering kernels from all previous historical events (i.e., all *t < t*_*i*_) and the background rate *µ*_*c*_. Therefore, we can consider the probability of case *i* caused by case *j, p*_*c*_(*i, j*), as the contribution of primary infection *j* in the event rate at time *t*_*i*_, i.e., *λ*_*c*_(*t*_*i*_), and *p*_*c*_(*i, i*) can be seen as the contribution of the background rate.

#### 2.3.2. Maximization step

We then maximize the complete data log-likelihood with respect to the model parameters, conditioned on the estimated branching structure *p*_*c*_(*i, j*). During estimation we do not include event pairs (*i, j*) when *j* is within Ψ = 14 days ^3^ of the last day of the dataset, as the offspring events *i* have not yet been realized and the inclusion of these incomplete data biases parameter estimates.

Given the latent variable *p*_*c*_(*i, j*), the maximization of Equation 6 can be decoupled into three independent optimization problems. Starting with the coefficient *θ* from Poisson regression, the maximization of likelihood function can be rewritten as the following:

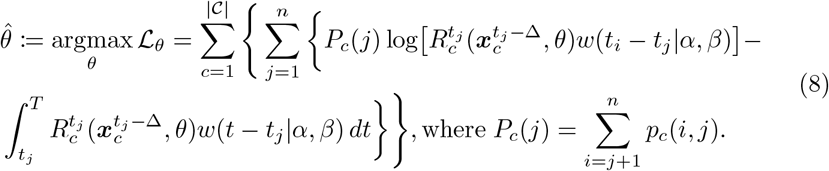

Because the last Ψ days are removed from the dataset and we assume that all possible offspring pairs (i, j) have been observed, we can therefore approximate the integrals for the inter-infection time *w*(*t*) in Equation 6 as is done in [28] by noting that 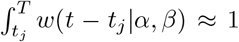. The optimization problem is therefore a Poisson regression, where we regress the observations 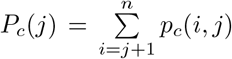 against the covariates 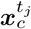.

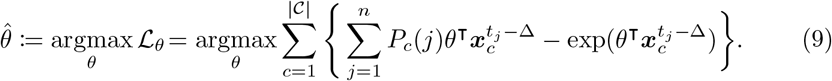

The same optimization strategy can be applied on the shape and scale parameters, *α* and *β*. The optimization problem can then be solved as a weighted maximum likelihood estimation for the Weibull shape and scale parameters:

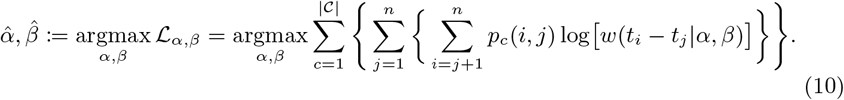

where *p*_*c*_(*i, j*) is the weight of each inter-infection time observation *t*_*i*_ *− t*_*j*_. Third, the background rate *µ*_*c*_ is determined analytically:

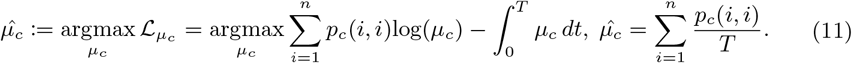

Pseudo code for the EM algorithm is presented in the Algorithm 1.

##### Algorithm 1 EM algorithm optimization

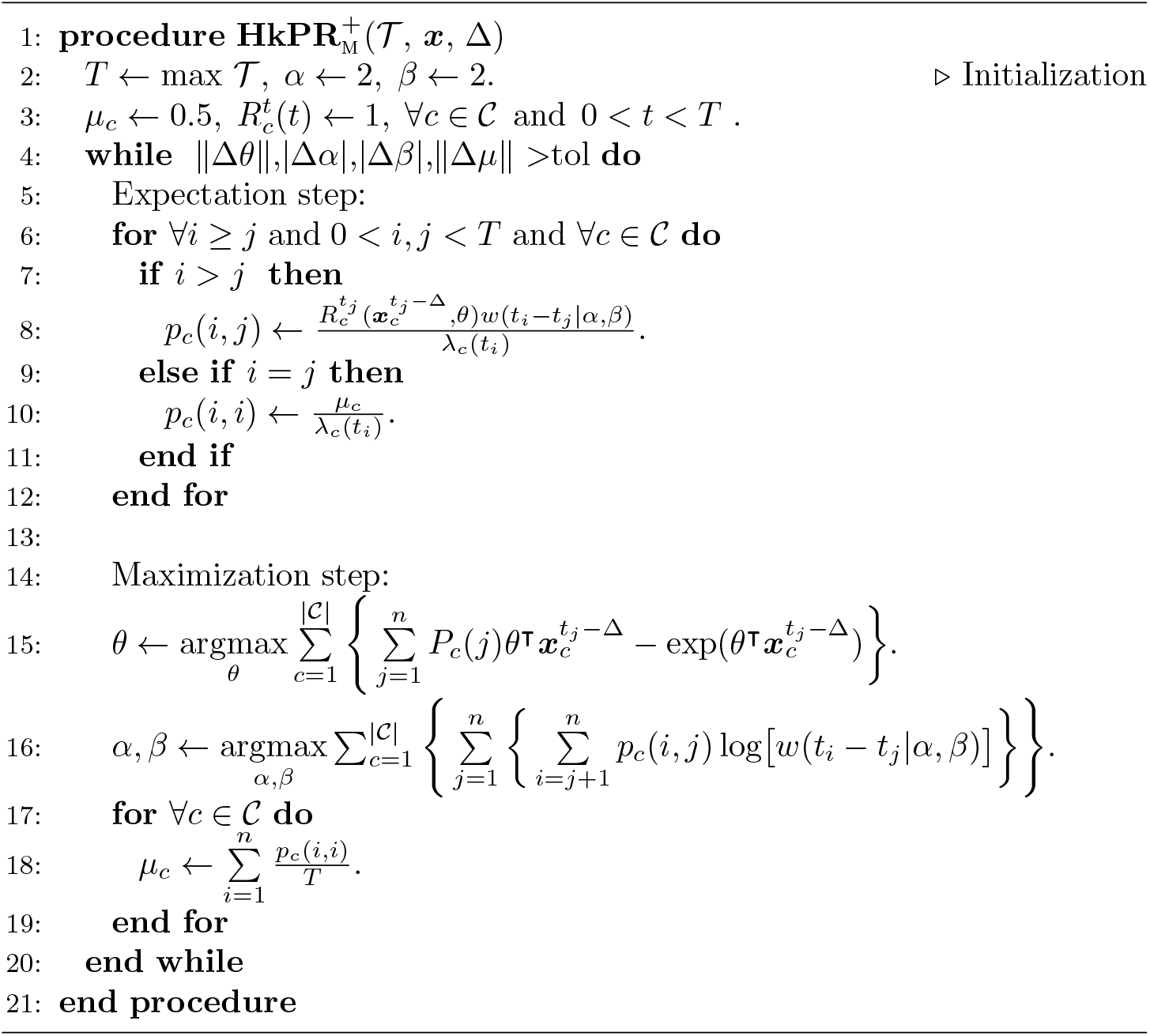

We note that the EM algorithm of the Hawkes process is also connected to the dynamic reproduction number estimator of Wallinga and Teunis [29], as the latter can be viewed as a 1-iteration EM algorithm where a histogram estimator is used for 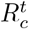 with initial guess 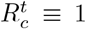. More details are discussed in the following section.

### 2.4. Connection of EM algorithm for Hawkes Process and dynamic R estimator of Wallinga and Teunis

Here we make the connection between the EM algorithm for the Hawkes process and the popular dynamic reproduction number estimator of Wallinga and Teunis [30, 29, 23]. The dynamic *R* estimator of Wallinga and Teunis is constructed as follows. The probability that individual *i* at time *t*_*i*_ was infected by individual *j* at time *t*_*j*_ is defined to be,

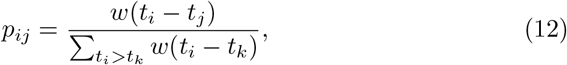

where the distribution of inter-infection times *w*(*t*_*i*_ *− t*_*j*_) is typically modeled as Weibull, Gamma, or log-normal [23]. The expected total number of individuals that *j* infects is then given by:

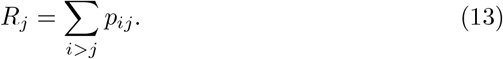

Wallinga and Teunis then obtain an estimate of the dynamic reproduction number *R*(*t*) by averaging *R*_*j*_ over all observed cases *j* where the time of infection *t*_*j*_ occurred on day *t*:

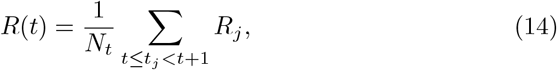

(here *N*_*t*_ is the number of observed infections on day *t*).

On the other hand, for the Hawkes process the intensity (rate) of infections is modeled as

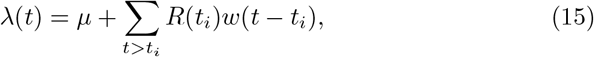

where *w*(*t*) and *R*(*t*) are the inter-infection time distribution and dynamic reproduction number respectively. Rather than modeling *R*(*t*) as dependent on mobility, we can instead model *R*(*t*) as a piece-wise constant function:

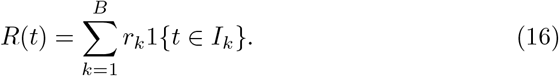

Here the *I*_*k*_ are intervals discretizing time, *B* is the number of such intervals, and *r*_*k*_ is the estimated reproduction rate in interval *k*.

Given initial guesses for the model parameters and *r*_*k*_, the EM algorithm for the Hawkes process iteratively updates the parameters and branching probabilities by alternating between the

**E-step update**:

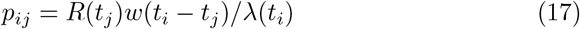

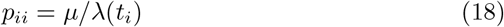

and **M-step update:**

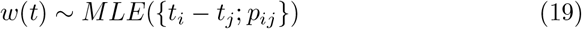

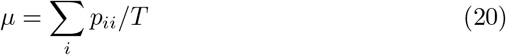

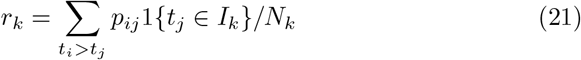

where *T* is the total length of the observation period, *N*_*k*_ is the total number of events in interval *k*, and the *w*(*t*) is estimated via weighted MLE (for either a Gamma, Weibull or log-normal) using the inter-event times as observations and branching probabilities as weights.

Finally, we can compare Equation 17 to Equation 12. The dynamic *R*(*t*) estimator in Equation 12 is what you obtain with 1 step of the EM algorithm in Equation 17 with initial guess *R*(*t*) *≡* 1, *µ* = 0 and 1 day chosen as the bin width for the histogram estimator.

### 2.5. Hawkes process forecasting

Finally, we forecast future events using the branching process representation of the Hawkes process. In particular, for each event in the history of the process we simulate a Poisson random variable with mean 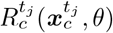 representing the number of secondary infections caused by event *j*. For each of these infections we simulate the time of infection by drawing inter-event times from the estimated Weibull distribution. Events falling in the future (past the forecasting date) are then used to update the forecasted intensity through Equation 1. We simulate multiple realizations of this process to estimate a mean intensity forecast along with confidence intervals.

## 3. Experiments and Results

In this section we first provide details on the datasets and baseline models used in our experiments. We then discuss the experimental results of several COVID-19 retrospective forecasting tasks at the U.S. county level. The source code and dataset are included in the supplemental material and are available online in a anonymous repository, respectively ^4^.

### 3.1. Datasets

#### 3.1.1. Covid-19 daily cases and deaths reported by The New York Times

The New York Times (NYT) [31] ^5^ releases a daily report of the cumulative numbers of COVID-19 cases in the United States at the county level and over time. While NYT data closely tracks data aggregated by a project at Johns Hopkins University [32], NYT county level reporting started earlier and is therefore used in this study. In total, there are 3, 217 counties with cases and/or deaths in the dataset. The time series data are compiled from state and local government health departments. In order to have sufficient data for statistical inference, we select the counties with confirmed cases greater than and equal to 10 (denoted by *𝒟*_conf_) and the counties with at least 1 death (denoted by *𝒟*_death_) by 11/10/2020 when the dataset is curated. In total, there are 2, 824 and 2, 545 counties in these two datasets. Parameter sharing may improve models in counties with less data through variance reduction, but can potentially bias estimates in more populated counties with more cases.

We therefore assess model performance over different subsets of counties grouped by case volume. We first rank counties by the number of confirmed cases and deaths by the cut-off date, 11/30/2020, and we then evaluate forecasting accuracy on the top-10% of counties (denoted by 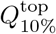), the top-25% counties (denoted by 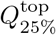), and counties between the top-25% and top-50% quantiles (denoted by 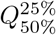).

In Figure 3a and 3b, we present the distribution of the cumulative confirmed cases and deaths at three different quantiles up to the cute-off date 11/30/2020. As the counties at the top-50% have more than 1,000 confirmed cases and 10 deaths, some urban counties, mostly at the top-10%, had already surpassed 10,000 confirmed cases and accumulated more than 300 deaths. In Figure 4a and 4b, we show the daily reported confirmed cases and deaths of top-3 counties in 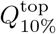 and 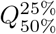 from *𝒟*_conf_ and *𝒟*_death_, respectively. Given different demographics and different COVID-19 regulations, each state went through different phases. For example, while Cook, IL seemed to contain the first spike after May, the confirmed cases in Los Angeles, CA seem steadily increase and only slow down after July. The daily death toll of Maricopa, AZ only hit its record high only after August unlike Los Angeles, CA, which had already had their first wave in terms of deaths in April. Overall, the deaths are increasing as the U.S. heads into the winter months. Such differences in infection rates suggest that different public health and social measures may need to be tailored county by county. Therefore, the proposed county-level forecasting model may aid local government policymakers in understanding the demographic and mobility factors that play a role in local reproduction of the virus.

**Figure 3:**
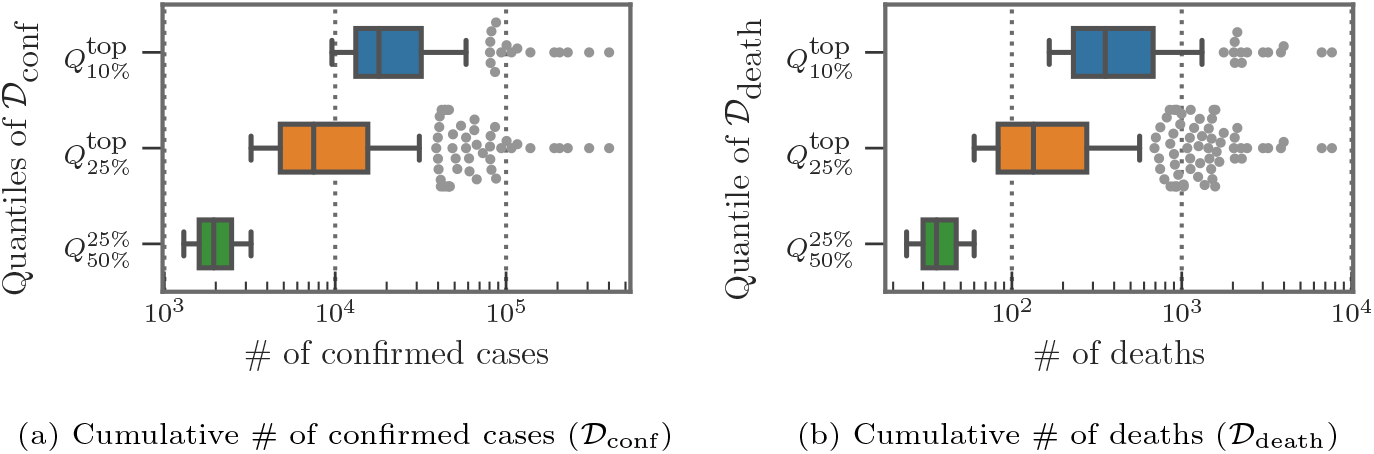
Distribution of cumulative cases reported at 11/30/2020 at different quantiles

**Figure 4:**
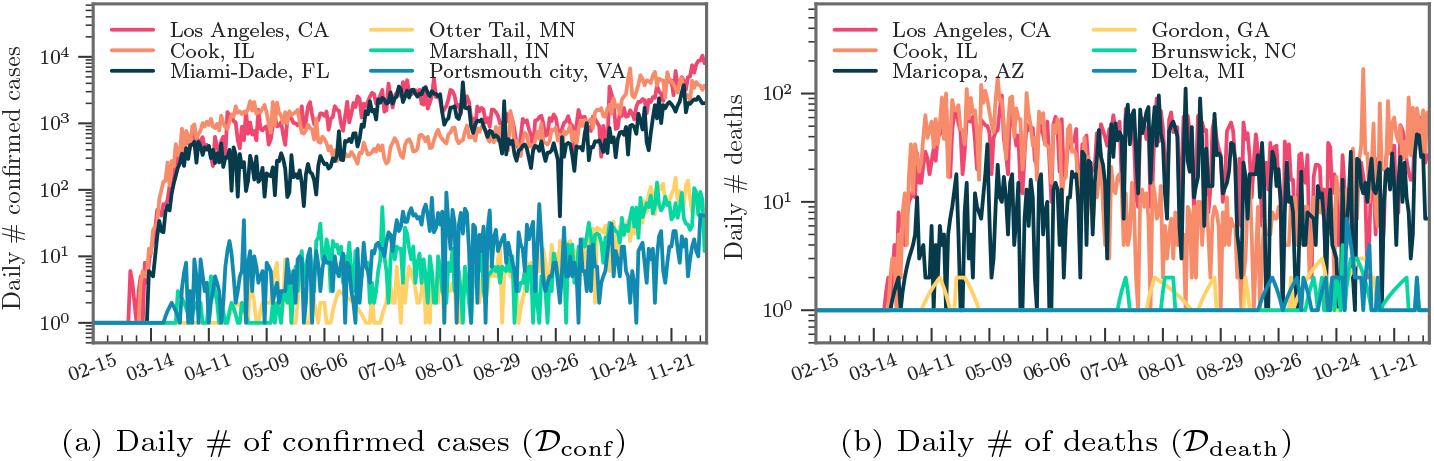
Example of the daily # of confirmed cases/deaths

#### 3.1.2. Google mobility index reports

We use Google daily mobility index reports at the county level [33] to estimate a dynamic reproduction number that tracks changes in movement patterns due to stay at home orders (and their staged removal). In total, there are 6 mobility types, including grocery & pharmacy, parks, transit stations, retail & recreation, residential and workplaces. Mobility indices for each category and county are calculated with respect to a baseline value for that day of the week ^6^. We drop “workplace” mobility from our analysis due to high collinearity with “residential” mobility. Some mobility data are missing when data is sparse for a given date. To deal with missing values, we adopt multivariate feature imputation ^7^, which estimates each missing mobility entry as a function of other mobility types on the same day in the same county. We show some heatmaps of mobility patterns across counties and time in the Figure 5, where a major change can be observed coinciding with stay at home orders (the first state-wide stay-at-home order was issued at 03/21/2020). Also, the reopening phase in most of the counties can be seen after May. For counties hit by COVID-19 the most (i.e., those in the top-10 %), we can also observe some strict regulations in the “Retail and recreation” areas and better compliance with stay-at-home orders based on high mobility in “Residential” area.

**Figure 5:**
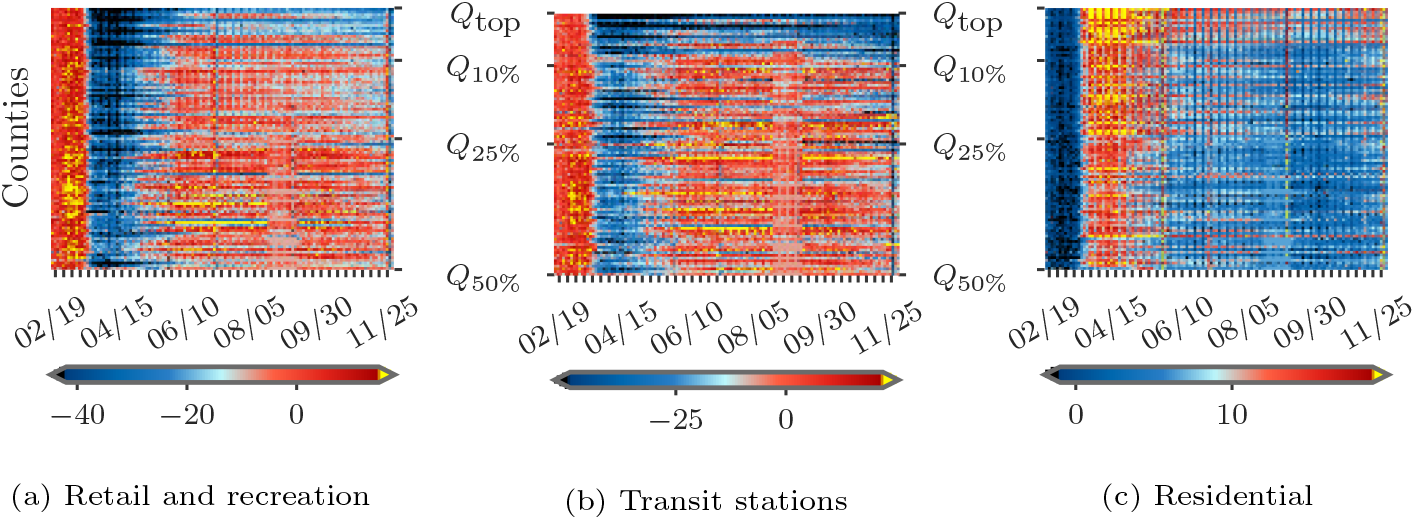
Heat map of mobility indices across counties in *𝒟*_conf_ and over time.

#### 3.1.3. County-level demographic covariates

We incorporate spatial demographic features that may be predictive of symptomatic cases of COVID-19 (which are more likely to result in testing and mortality). The dataset is available in a curated form [20] and is derived from CDC and census datasets. The data is at the county level and includes population, median age, number of hospitals and ICU beds, percentage of smokers and diabetes, and heart disease mortality.

In Figure 6, we present two examples of spatial demographic features at the county-level used to model variations in the reproduction number. In Figure 6a we observe that both the east and west coasts of the United States are more densely populated compared to midwestern and western regions. Diabetes percentage (shown in Figure 6b), on the other hand, is mostly higher in southern regions of the U.S.

**Figure 6:**
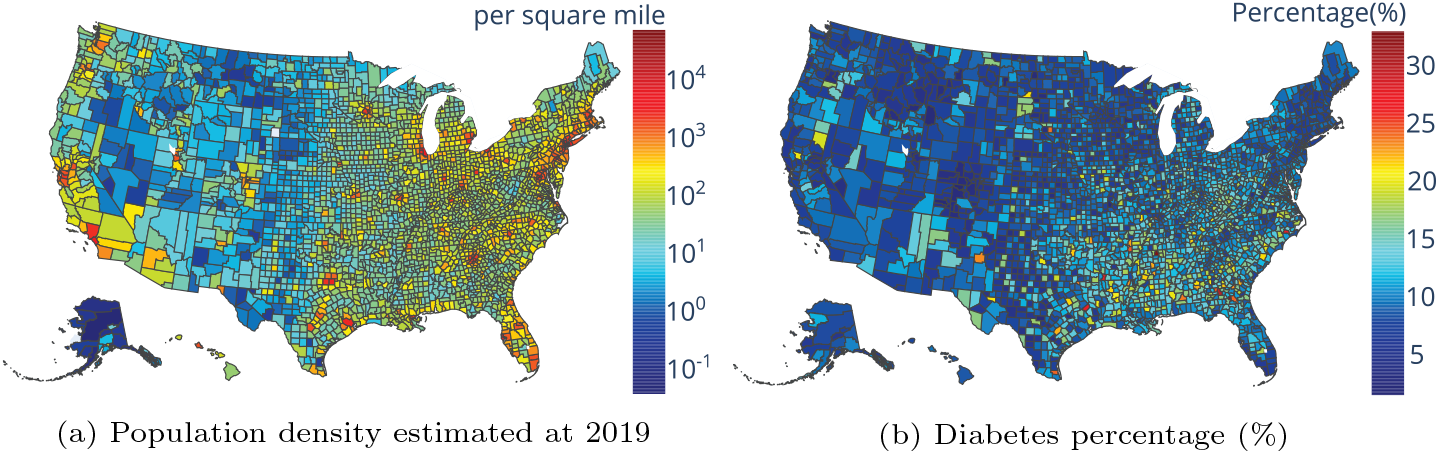
Examples of spatial demographic and health features at the county-level.

### 3.2. Baseline models and experimental setup for retrospective forecasting comparison

We compare the Hawkes process model in Equation 1 with several benchmarks including an SEIR model used in a pandemic tracking dashboard ^8^ out of Columbia University [2] (denoted by **PROJ**), an geospatial SEIR Model from the Johns Hopkins University Applied Physics Lab [4] (denoted by **BUCKY**), and an ensemble model with linear and exponential predictors from University of California, Berkeley [20] (denoted by **CLEP**). Note that all three benchmarks are tested directly from the released source code and we follow the same experimental protocol as for our proposed model. A simplified Hawkes process, denoted by **Hawkes**, where the reproduction number is held constant is used for comparison to demonstrate the effectiveness of tracking the reproduction number dynamically. We also compare our full Hawkes process model, denoted by 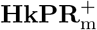, to a Hawkes process, **HkPR**_m_, with only mobility features to determine the marginal improvement of adding demographics.

We backtest the six competing models on the 𝒟_conf_ and 𝒟_death_ datasets using the “walk-forward” validation approach. In particular, for 7-day forecasts we first train the models based on cases and deaths before the first cut-off date, 04/15/2020, and then forecast through 04/21/2020. We then slide the forecasting window, training on data before 4/22/2020 and forecasting from 04/22/2020 to 04/28/2020. We repeat this process until the final date of 05/19/2020 (a similar approach is used for 14 and 28 day forecasts). The multivariate imputation models are also trained in the same walk forward fashion to avoid possible data leakage. The hyper-parameter of the lag parameter Δ ranges from 7, 14, 21, and 28 days in our experiments.

We evaluate the models according to mean absolute error, `MAE`, averaged across counties and forecasting windows of the same length, along with percentage error, `PE`. Mean absolute error (`MAE`) and the percentage error (`PE`) are calculated as follows:

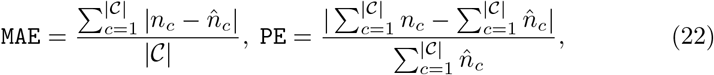

where 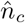, and 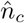 are the number of reported events and predicted events, respectively. We also compare the ranking quality of the competing models using Normalized Discounted Cumulative Gain (`NDCG`) [34], which can be used to evaluate the power of recommendations for counties with potential COVID-19 spikes in the near future.

### 3.3. Experimental results

In Table 1 and Table 2, we present the experimental results for 7, 14, and 28 days window forecasts of `MAE` for all models applied to both confirmed cases (*𝒟*_conf_) and deaths (*𝒟*_death_), and in Table 3 and Table 4, we report the results for `PE`. In terms of `MAE` and `PE`, both of our proposed models, **HkPR**_m_ and 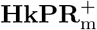, outperform the benchmarks, **PROJ** and **CLEP**, by a large margin in all three forecasting periods and across quantile subsets of the data.The improvements of `MAE` and `PE` can also be seen in the simplistic baseline Hawkes process, **Hawkes**. This suggests that the Hawkes process approach has a good potential on modeling infectious disease due to the self-exciting properties that lie in the COVID-19 cases.

**Table 1:** `MAE` on predicted confirmed cases *𝒟*_conf_

| Model | 7-days |  |  | 14-days |  |  | 28-days |  |  |
| --- | --- | --- | --- | --- | --- | --- | --- | --- | --- |
| | $Q_{10\%}^{\text{top}}$ | $Q_{25\%}^{\text{top}}$ | $Q_{50\%}^{25\%}$ | $Q_{10\%}^{\text{top}}$ | $Q_{25\%}^{\text{top}}$ | $Q_{50\%}^{25\%}$ | $Q_{10\%}^{\text{top}}$ | $Q_{25\%}^{\text{top}}$ | $Q_{50\%}^{25\%}$ |
| **PROJ** | 809.40 | 415.88 | 55.30 | 1664.90 | 857.93 | 117.86 | 3432.57 | 1779.56 | 252.43 |
| **CLEP** | 238.30 | 134.83 | 33.86 | 585.81 | 324.32 | 88.87 | 1963.52 | 1090.36 | 207.81 |
| **BUCKY** | 404.49 | 212.80 | 37.45 | 883.69 | 459.77 | 89.85 | 2085.88 | 1116.91 | 229.33 |
| **Hawkes** | 224.35 | 120.61 | 24.02 | 569.49 | 300.06 | 55.45 | 1803.63 | 935.83 | 165.92 |
| **HkPR<sub>m</sub>** | 211.59 | **114.34** | 22.44 | **519.00** | **271.86** | **49.83** | **1573.58** | **835.59** | 136.60 |
| **HkPR<sub>m</sub><sup>+</sup>** | **210.72** | 114.69 | **22.38** | 522.92 | 276.28 | 49.86 | 1611.48 | 893.65 | **132.79** |

The best performance is marked in **bold**.

**Table 2:** `MAE` on predicted confirmed cases *𝒟*_death_

| Model | 7-days |  |  | 14-days |  |  | 28-days |  |  |
| --- | --- | --- | --- | --- | --- | --- | --- | --- | --- |
| | $Q_{10\%}^{\text{top}}$ | $Q_{25\%}^{\text{top}}$ | $Q_{50\%}^{25\%}$ | $Q_{10\%}^{\text{top}}$ | $Q_{25\%}^{\text{top}}$ | $Q_{50\%}^{25\%}$ | $Q_{10\%}^{\text{top}}$ | $Q_{25\%}^{\text{top}}$ | $Q_{50\%}^{25\%}$ |
| **PROJ** | 15.56 | 7.89 | 1.12 | 30.66 | 15.55 | 2.20 | 56.85 | 29.37 | 4.41 |
| **CLEP** | 10.96 | 5.83 | 1.16 | 19.10 | 10.58 | 2.31 | 78.17 | 42.99 | 8.56 |
| **BUCKY** | 8.23 | 4.55 | **1.00** | 16.07 | 8.70 | 1.83 | **29.55** | **16.56** | 3.98 |
| **Hawkes** | 8.49 | 4.59 | 1.04 | 17.38 | 9.18 | 1.98 | 47.13 | 24.29 | 4.32 |
| **HkPR<sub>m</sub>** | **7.19** | **4.07** | 1.01 | **13.40** | 7.55 | 1.78 | 33.30 | 18.23 | 3.74 |
| **HkPR<sub>m</sub><sup>+</sup>** | 7.24 | 4.07 | 1.01 | 13.68 | **7.53** | **1.77** | 35.99 | 19.18 | **3.60** |

The best performance is marked in **bold**.

**Table 3:** `PE` on predicted confirmed cases *𝒟*_conf_

| Model | 7-days |  |  | 14-days |  |  | 28-days |  |  |
| --- | --- | --- | --- | --- | --- | --- | --- | --- | --- |
| | $Q_{10\%}^{\text{top}}$ | $Q_{25\%}^{\text{top}}$ | $Q_{50\%}^{25\%}$ | $Q_{10\%}^{\text{top}}$ | $Q_{25\%}^{\text{top}}$ | $Q_{50\%}^{25\%}$ | $Q_{10\%}^{\text{top}}$ | $Q_{25\%}^{\text{top}}$ | $Q_{50\%}^{25\%}$ |
| **PROJ** | 91.00 | 90.40 | 84.07 | 94.41 | 94.63 | 92.83 | 95.76 | 96.17 | 96.94 |
| **CLEP** | 10.23 | 12.08 | 41.09 | 24.60 | 29.42 | 90.24 | 41.19 | 47.74 | 515.17 |
| **BUCKY** | 19.61 | 20.12 | 35.58 | 28.05 | 27.90 | 69.61 | 47.91 | 49.24 | 97.86 |
| **Hawkes** | 11.52 | 11.31 | 14.36 | 17.33 | 17.02 | 19.60 | 41.25 | 40.06 | **39.11** |
| **HkPR<sub>m</sub>** | 11.72 | 10.75 | 15.44 | **13.92** | 15.10 | **15.08** | **38.77** | 38.20 | 46.38 |
| **HkPR<sub>m</sub><sup>+</sup>** | **10.16** | **10.35** | **12.95** | 15.30 | **13.45** | 16.91 | 41.96 | **33.33** | 41.31 |

The best performance is marked in **bold**.

**Table 4:** `PE` on predicted confirmed cases *𝒟*_death_

| Model | 7-days |  |  | 14-days |  |  | 28-days |  |  |
| --- | --- | --- | --- | --- | --- | --- | --- | --- | --- |
| | $Q_{10\%}^{\text{top}}$ | $Q_{25\%}^{\text{top}}$ | $Q_{50\%}^{25\%}$ | $Q_{10\%}^{\text{top}}$ | $Q_{25\%}^{\text{top}}$ | $Q_{50\%}^{25\%}$ | $Q_{10\%}^{\text{top}}$ | $Q_{25\%}^{\text{top}}$ | $Q_{50\%}^{25\%}$ |
| **PROJ** | 72.56 | 72.25 | 45.10 | 81.93 | 81.96 | 73.62 | 90.21 | 90.53 | 87.16 |
| **CLEP** | 23.39 | 26.35 | 18.05 | 19.23 | 19.27 | 24.74 | 73.77 | 78.90 | 173.88 |
| **BUCKY** | 17.64 | 16.08 | 14.71 | **15.63** | **13.93** | 20.22 | **15.36** | **13.20** | 36.19 |
| **Hawkes** | 17.97 | 16.99 | 15.81 | 20.03 | 20.71 | 16.17 | 48.79 | 44.15 | 28.17 |
| **HkPR<sub>m</sub>** | **16.77** | **15.59** | **13.80** | 20.40 | 17.38 | **13.72** | 33.03 | 52.51 | 22.04 |
| **HkPR<sub>m</sub><sup>+</sup>** | 17.53 | 16.92 | 14.05 | 18.18 | 15.23 | 16.93 | 38.31 | 44.78 | **17.66** |

The best performance is marked in **bold**.

We found that adding mobility indices improves **Hawkes**, where forecasting accuracy of **HkPR**_m_ also increases across the subsets and all forecasting window. For example, the improvements on `MAE` over **Hawkes** can go up to 13%, 11%, and 18% for 28 days forecast when **HkPR**_m_ is applied to 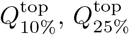, and 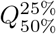 in *𝒟*_conf_, respectively. Similar decrease on `MAE` can be observed when **HkPR**_m_ is applied to three quantile subsets in _death_, where **HkPR**_m_ outperforms **Hawkes** by 29%, 25%, and 13% in `MAE`, respectively. In terms of `PE`, **HkPR**_m_ stays ahead of **Hawkes** with only one exception at 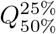 of 𝒟_death_ in 28 days forecasting. This shows that by modeling the reproduction number through daily mobility indices we can enhance the forecasting accuracy and obtain more precise estimation on the spikes in the future.

By adding demographic features, we can marginally boost the `MAE` and `PE` of 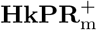 over **HkPR**_m_ in some cases. In general, the variation, 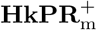, also shows similar improvements over the benchmarks. In particular, 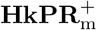 has the best PE enhancement over **HkPR**_m_ at 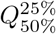 in *𝒟*_death_ for 28 days forecast, which is 20%. This demonstrates that the major forecasting power comes from the joint modeling of mobility indices in the reproduction number while the choices of the background rate and inter-infection distribution may only play a minor part.

Moreover, we notice that model **BUCKY** is a competitive baseline in *𝒟*_death_ where it has better accuracy in a few cases, such as `MAE` and `PE` 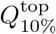 and 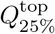 for 28 days forecast. Possible explanation for its advantage could be the CDC-recommended parameters that has been introduced to aid the model training especially for recovery and deaths compartments in its SEIR model. Those parameters include case fatality ratio, case hospitalization ratio, time between death and reporting, etc. However, introducing such pre-trained parameters from CDC may not be practical in real-time forecasting and may potentially bring in the data leakage issue.

In Table 5 and Table 6, we present the `NDCG` results for the ranking evaluation. Generally, the proposed models **HawkPR** have a better `NDCG` performance when applied to confirmed cases for most of the quantile subsets. In terms of `NDCG` on the 𝒟_death_ dataset, the baseline Hawkes process, **Hawkes**, performs better in some cases but proposed method consistently comes in second for most of the forecasting window. By generating rankings with good qualities, **HawkPR** can serve as a recommender system for the hotspot counties and the public health policymakers can tailor strategies specifically for each region to contain the virus. We also note that in our model, we are estimating inter-event distributions of observed cases (ignoring asymptomatic cases) and therefore these are observed or “effective” inter-event distributions, rather than true inter-infection distributions based on longitudinal data (we will clarify the language in the revision). We believe this approach is justified by the performance of the model in forecasting observed cases (and this approach is taken in other applications, like seismology where some earthquakes are not observed).

**Table 5:** `NDCG` on predicted confirmed cases *𝒟*_conf_

| Model | 7-days |  |  | 14-days |  |  | 28-days |  |  |
| --- | --- | --- | --- | --- | --- | --- | --- | --- | --- |
| | $Q_{10\%}^{\text{top}}$ | $Q_{25\%}^{\text{top}}$ | $Q_{50\%}^{25\%}$ | $Q_{10\%}^{\text{top}}$ | $Q_{25\%}^{\text{top}}$ | $Q_{50\%}^{25\%}$ | $Q_{10\%}^{\text{top}}$ | $Q_{25\%}^{\text{top}}$ | $Q_{50\%}^{25\%}$ |
| **PROJ** | 0.7225 | 0.7039 | 0.8232 | 0.6946 | 0.6793 | 0.8412 | 0.6696 | 0.6614 | 0.8589 |
| **CLEP** | 0.9626 | 0.9526 | 0.8620 | 0.9418 | 0.9402 | 0.8704 | 0.9015 | 0.8843 | 0.8739 |
| **BUCKY** | 0.9283 | 0.9279 | 0.8600 | 0.9269 | 0.9216 | 0.8768 | 0.9013 | 0.8957 | 0.8813 |
| **Hawkes** | **0.9738** | 0.9757 | **0.8680** | 0.9697 | 0.9704 | 0.8926 | 0.9414 | 0.9419 | 0.8879 |
| **HkPR<sub>m</sub>** | 0.9706 | 0.9728 | 0.8673 | 0.9715 | 0.9755 | **0.8956** | **0.9502** | **0.9521** | **0.8958** |
| **HkPR<sub>m</sub><sup>+</sup>** | 0.9734 | **0.9759** | 0.8672 | **0.9752** | **0.9758** | 0.8932 | 0.9493 | 0.9503 | 0.8918 |

The best performance is marked in **bold**.

**Table 6:** `NDCG` on predicted confirmed cases *𝒟*_death_

| Model | 7-days |  |  | 14-days |  |  | 28-days |  |  |
| --- | --- | --- | --- | --- | --- | --- | --- | --- | --- |
| | $Q_{10\%}^{\text{top}}$ | $Q_{25\%}^{\text{top}}$ | $Q_{50\%}^{25\%}$ | $Q_{10\%}^{\text{top}}$ | $Q_{25\%}^{\text{top}}$ | $Q_{50\%}^{25\%}$ | $Q_{10\%}^{\text{top}}$ | $Q_{25\%}^{\text{top}}$ | $Q_{50\%}^{25\%}$ |
| **PROJ** | 0.6746 | 0.6607 | 0.7057 | 0.6823 | 0.6529 | 0.7666 | 0.7003 | 0.6837 | 0.8050 |
| **CLEP** | 0.9126 | 0.9010 | 0.7451 | 0.9095 | 0.8945 | 0.7849 | 0.8696 | 0.8240 | 0.8043 |
| **BUCKY** | 0.9161 | 0.9160 | 0.7301 | 0.9217 | 0.9222 | 0.7797 | 0.9074 | 0.9095 | **0.8278** |
| **Hawkes** | **0.9506** | **0.9493** | 0.7548 | 0.9475 | 0.9469 | **0.8011** | 0.9293 | 0.9297 | 0.8212 |
| **HkPR<sub>m</sub>** | 0.9491 | 0.9476 | **0.7598** | 0.9446 | 0.9457 | 0.8007 | 0.9299 | 0.9315 | 0.8195 |
| **HkPR<sub>m</sub><sup>+</sup>** | 0.9504 | 0.9474 | 0.7597 | **0.9502** | **0.9514** | 0.7963 | **0.9368** | **0.9372** | 0.8176 |

The best performance is marked in **bold**.

#### 3.3.1. Importance of covariates

In Table 7 and Table 8, we show the dynamic reproduction number coefficients of 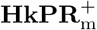 estimated from the Poisson regression component (Equation 2) when applied to 𝒟_conf_ and 𝒟_death_, respectively. The absolute value of the coefficients indicates the magnitude of the correlation between the reproduction number and the features. With the exception of population estimation in 𝒟_death_, the coefficients of all variables are statistically significant at the 10^*−*7^ level or below. The dynamic reproduction number is positively correlated with “Retail and recreation” while negatively correlated with “Residential”, meaning that as mobility shifted away from commercial areas towards residences, the reproduction number decreased. In terms of spatial covariates, the reproduction number is positively correlated with “Population density” and “# of ICU beds.” This suggests that the regions hit the hardest by COVID-19 are mostly urban areas, where most of intensive treatment units are situated. The reproduction number is also negatively correlated with percent of the population with” Diabetes” and “Heart disease mortality rate.” Several possible explanations for this observation include high-risk individuals are being more cautious or that they tend to live in areas with less cases, potentially with less population.

**Table 7:**
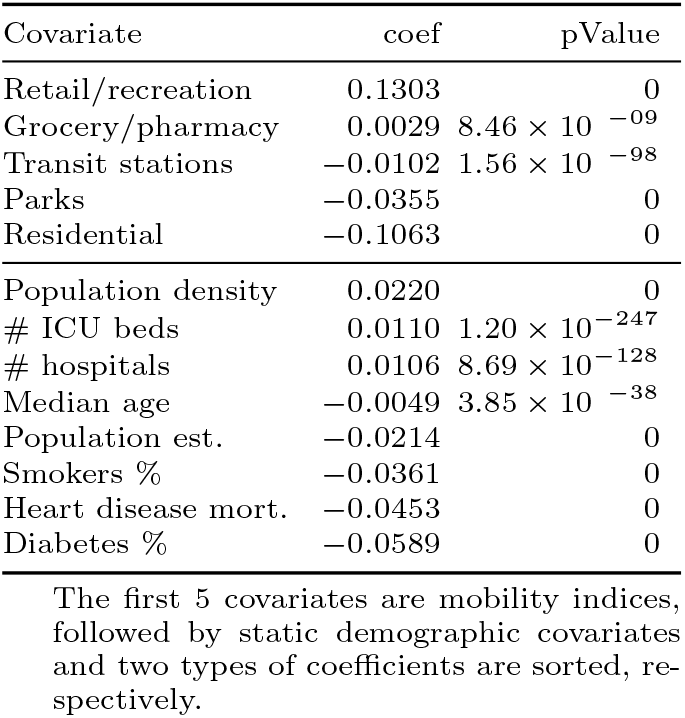
Model coefficients (*𝒟*_conf_)

| Covariate | coef | pValue |
| --- | --- | --- |
| Retail/recreation | 0.1303 | 0 |
| Grocery/pharmacy | 0.0029 | $8.46 \times 10^{-09}$ |
| Transit stations | -0.0102 | $1.56 \times 10^{-98}$ |
| Parks | -0.0355 | 0 |
| Residential | -0.1063 | 0 |
| Population density | 0.0220 | 0 |
| # ICU beds | 0.0110 | $1.20 \times 10^{-247}$ |
| # hospitals | 0.0106 | $8.69 \times 10^{-128}$ |
| Median age | -0.0049 | $3.85 \times 10^{-38}$ |
| Population est. | -0.0214 | 0 |
| Smokers % | -0.0361 | 0 |
| Heart disease mort. | -0.0453 | 0 |
| Diabetes % | -0.0589 | 0 |

The first 5 covariates are mobility indices, followed by static demographic covariates and two types of coefficients are sorted, respectively.

**Table 8:** Model coefficients (*𝒟*_death_)

| Covariate | coef | pValue |
| --- | --- | --- |
| Retail/recreation | 0.1047 | $4.15 \times 10^{-118}$ |
| Grocery/pharmacy | 0.0746 | $8.88 \times 10^{-111}$ |
| Transit stations | 0.0276 | $2.48 \times 10^{-11}$ |
| Residential | -0.0929 | $5.04 \times 10^{-212}$ |
| Parks | -0.1294 | 0 |
| # ICU beds | 0.0423 | $2.17 \times 10^{-64}$ |
| Population density | 0.0409 | 0 |
| Population est. | 0.0062 | $5.18 \times 10^{-2}$ |
| Median age | -0.0157 | $1.65 \times 10^{-7}$ |
| Heart disease mort. | -0.0250 | $1.29 \times 10^{-8}$ |
| # hospitals | -0.0423 | $6.54 \times 10^{-28}$ |
| Diabetes % | -0.1041 | $1.33 \times 10^{-86}$ |
| Smokers % | -0.1448 | $1.65 \times 10^{-279}$ |

The first 5 covariates are mobility indices, followed by static demographic covariates and two types of coefficients are sorted, respectively.

#### 3.3.2. COVID-19 forecasting and reproduce number analysis

In Figure 7, we present an example of 28 days projection made through 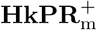 from 10/28/2020 - 11/25/2020 for both 𝒟_conf_ and 𝒟_death_. We can observe that 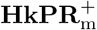 has very promising results in making projections, especially for the short term future, When the number of forecasting windows increases, the forecasting error increase as the task also being more difficult. Moreover, the narrow confidence interval calculated through 100 Hawkes processes simulations suggests that the the proposed model can make relatively stable forecasting. Lastly, based on the projections, as the number of confirmed cases soon would hit over 500,000 in the top counties including Los Angeles, CA, Cook, IL, and etc. It is imperative to have a robust framework to help governments to design strategies to combat COVID-19 or even more, prioritize vaccine distribution.

**Figure 7:**
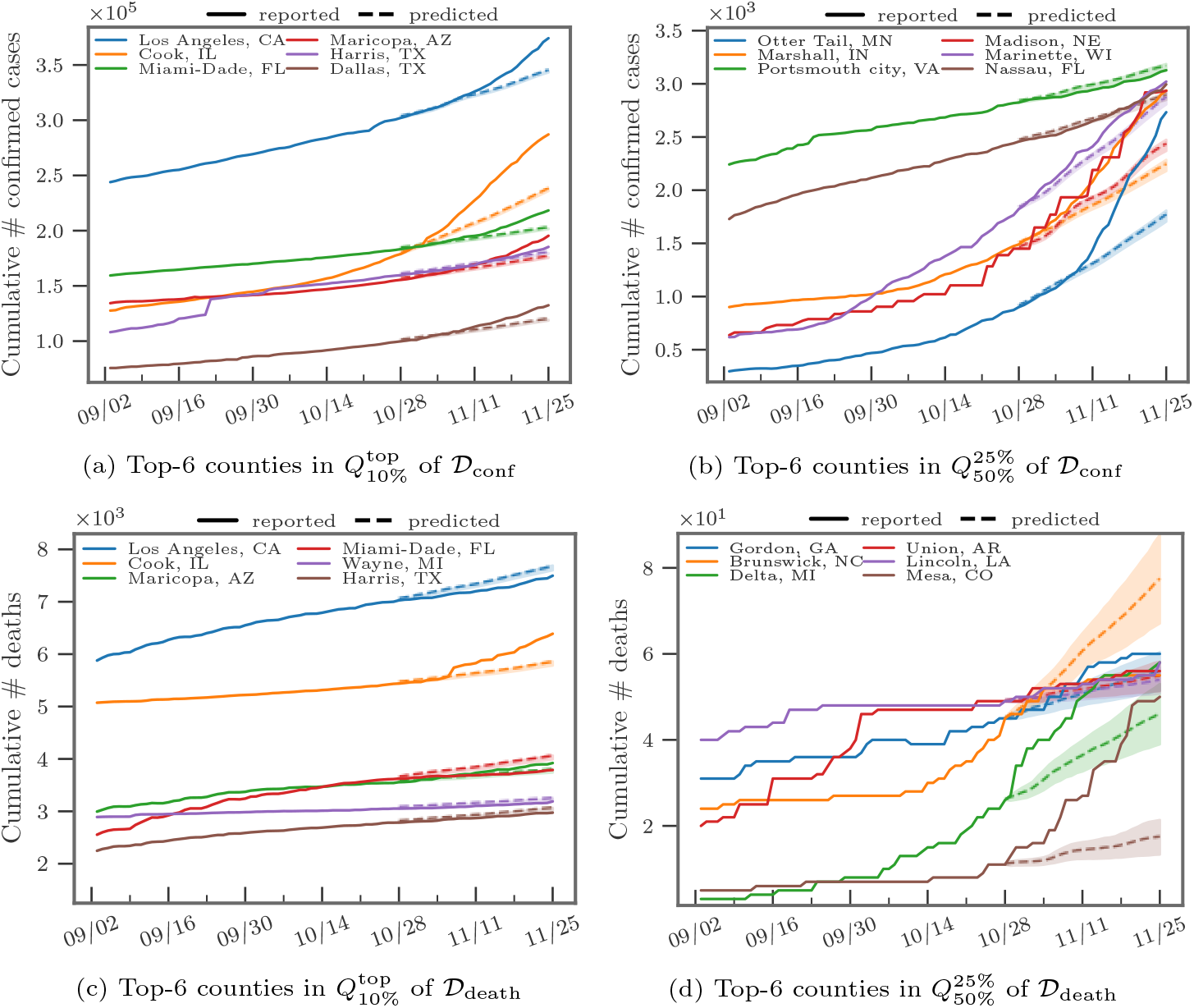
Forecasting for 28 days from 10/28/2020 - 11/25/2020

In Figure 8 and Figure 9, we find that the estimated dynamic reproduction number closely tracks lagged mobility, where the optimal lag parameter is determined as Δ = 14 days for *𝒟*_conf_ and Δ = 21 days for *𝒟*_death_. The top-2 counties in 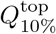 have estimated reproduction number initially above 2.5. After stay-at-home orders (around 04/11/2020), mobility in residential areas increased. On the other hand, mobility in retail and recreation decreased and the reproduction number fell to around 1, which explains why curves were relatively “flat” in many areas in the U.S. after the lockdown. However, as most of states reopened and lifted up the restrictions, the reproduction number increased after a large population resumed their daily routine, which can be also be observed by the increased mobility in retail and recreation after July. Lastly, to validate the reproduction numbers, we also compare our results to the ones estimated by Stanford University ^9^ and our estimation match to their findings, which are around 1.5-2.5 initially and 0.5-1.5 up to the beginning of December in 2020.

**Figure 8:**
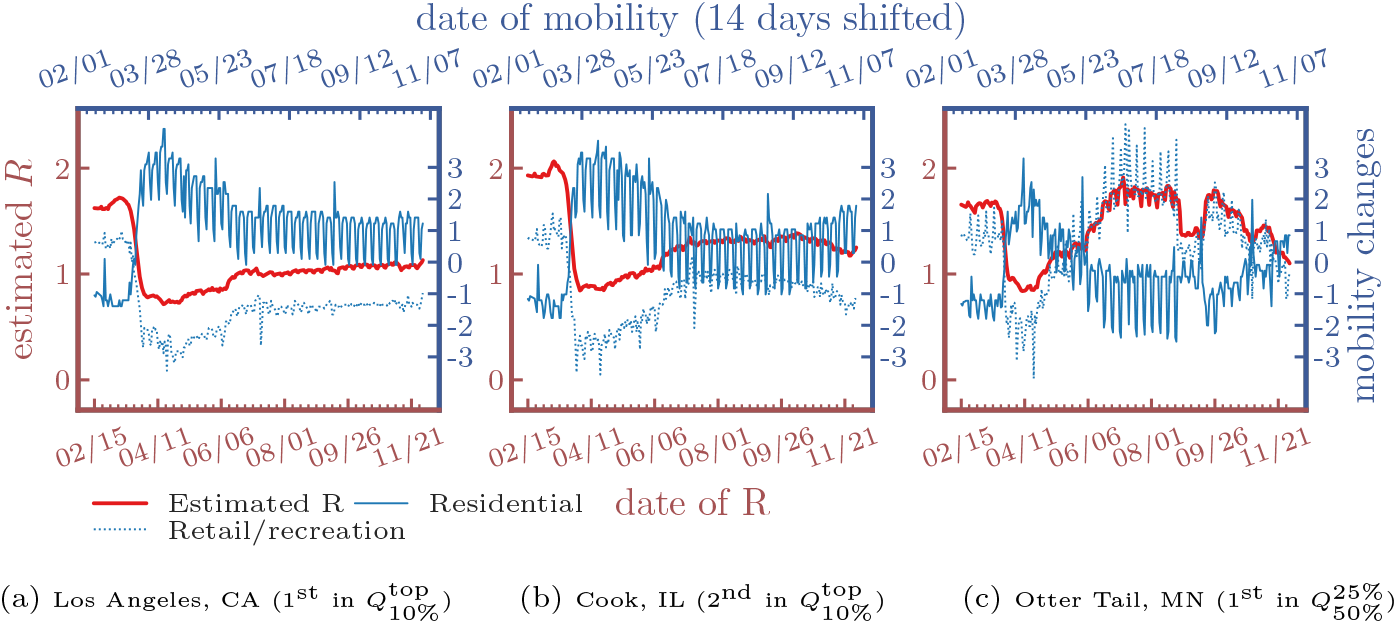
Estimated R of confirmed cases *𝒟*_conf_ and lagged mobility changes (Δ = 14 days)

**Figure 9:**
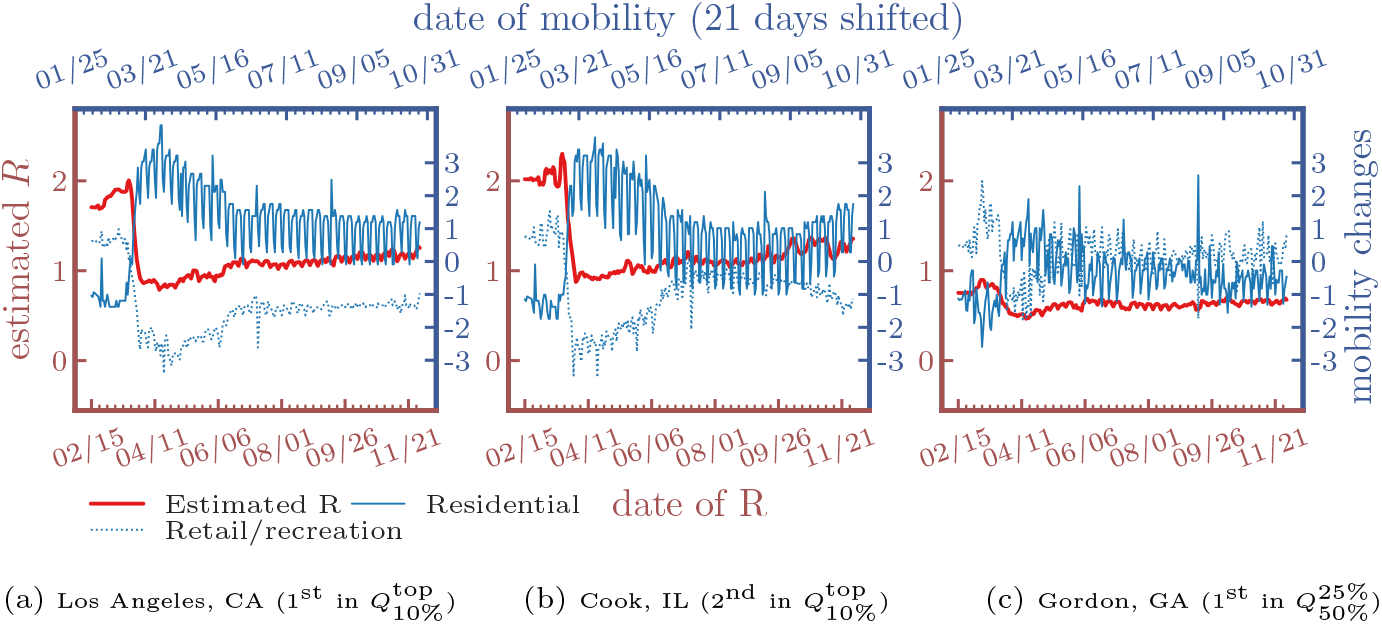
Estimated R of deaths *𝒟*_death_ and lagged mobility changes (Δ = 21 days)

## 4. Conclusion

We showed how Hawkes processes can be combined with spatio-temporal covariates to accurately model COVID-19 transmission and forecast future cases and deaths. The model is competitive with several benchmark models used to forecast the pandemic, achieving improved MAE and NDCG scores on a majority of the experiments we conducted. Our hope is that this work will encourage more research into Hawkes process models of disease spreading that incorporate more advanced features and machine learning principles. As vaccinations are rolled out across the U.S. (given recent FDA approval), local impacts on dynamic reproduction can be flexibly accommodated by our model and used to obtain more accurate and timely forecasts.

One potential direction for future research is extending the work here to neural network based point process models [10, 11]. These models may be able to capture more complicated relationships between mobility patterns, demographics, and transmission. The challenges of such an approach include the potential for over-fitting with added parameters and determining how best to realistically model transmission in a neural point process (analagous to the SIR-Hawkes process), which will be important if neural point processes are to be used in long-term forecasting.

Many of the preprints and models currently being released on academic archives and websites present a single model without model evaluation, goodness of fit analysis, or comparison to baselines. Here we believe the “common task framework” [35] could be beneficial in model selection and validation. The machine learning community can contribute to pandemic modeling efforts by performing careful benchmarking of methodologies, creating standardized datasets and tasks, and comparing competing models that come from different fields such as epidemiology, statistics, and machine learning.

## Data Availability

The source code and pruned dataset is available online.

https://github.com/chiangwe/HawkPR

## Footnotes

1 https://ai.facebook.com/blog/using-ai-to-help-health-experts-address-the-covid-19-pandemic

2 https://covid19forecasthub.org/community/

3 We choose Ψ = 14 as the incubation period for COVID-19 is thought to extend to 14 days given by the Clinical Care Guidance from the CDC: https://www.cdc.gov/coronavirus/2019-ncov/hcp/clinical-guidance-management-patients.html.

4 https://anonymous.4open.science/r/d425dcf9-3cfb-4f82-a08c-ee583ab36291/

5 https://github.com/nytimes/covid-19-data

6 The baseline is the median value, for the corresponding day of the week calculated during the 5-week period, 01/03/2020 to 02/06/2020.

7 https://scikit-learn.org/stable/modules/impute.html#multivariate-feature-imputation

8 https://covid19forecasthub.org/community/

9 https://web.stanford.edu/chadj/Covid/Dashboard.html

## References

[1] N. M. Ferguson, D. Laydon, G. Nedjati-Gilani, N. Imai, K. Ainslie, M. Baguelin, S. Bhatia, A. Boonyasiri, Z. Cucunubá, G. Cuomo-Dannenburg, et al., Impact of non-pharmaceutical interventions (npis) to reduce covid-19 mortality and healthcare demand. 2020, DOI 10 (2020) 77482.

[2] S. Pei, J. Shaman, Initial simulation of sars-cov2 spread and intervention effects in the continental us, medRxiv.

[3] M. Smith, K. Yourish, S. Almukhtar, K. Collins, D. Ivory, A. McCann, et al., Coronavirus in the us: Latest map and case count, The New York Times.

[4] M. Kinsey, K. Tallaksen, O. R.F., L. Asher, C. Costello, M. Kelbaugh, S. Wilson, Bucky model: a spatial seir model for simulating covid-19 at the county level. URL https://buckymodel.com/

[5] D. Zou, L. Wang, P. Xu, J. Chen, W. Zhang, Q. Gu, Epidemic model guided machine learning for covid-19 forecasts in the united states, medRxiv.

[6] I. Covid, C. J. Murray, et al., Forecasting covid-19 impact on hospital bed-days, icu-days, ventilator-days and deaths by us state in the next 4 months, medRxiv.

[7] N. P. Jewell, J. A. Lewnard, B. L. Jewell, Caution warranted: using the institute for health metrics and evaluation model for predicting the course of the covid-19 pandemic, Annals of Internal Medicine.

[8] M.-A. Rizoiu, S. Mishra, Q. Kong, M. Carman, L. Xie, Sir-hawkes: linking epidemic models and hawkes processes to model diffusions in finite populations, in: Proceedings of the 2018 World Wide Web Conference, 2018, pp. 419–428.

[9] K. Zhou, H. Zha, L. Song, Learning triggering kernels for multi-dimensional hawkes processes, in: International Conference on Machine Learning, 2013, pp. 1301–1309.

[10] H. Mei, J. M. Eisner, The neural hawkes process: A neurally self-modulating multivariate point process, in: Advances in Neural Information Processing Systems, 2017, pp. 6754–6764.

[11] T. Omi, K. Aihara, et al., Fully neural network based model for general temporal point processes, in: Advances in Neural Information Processing Systems, 2019, pp. 2120–2129.

[12] G. Mohler, M. D. Porter, Rotational grid, pai-maximizing crime forecasts, Statistical Analysis and Data Mining: The ASA Data Science Journal 11 (5) (2018) 227–236.

[13] H. Xu, H. Zha, A dirichlet mixture model of hawkes processes for event sequence clustering, in: Advances in Neural Information Processing Systems, 2017, pp. 1354–1363.

[14] N. Du, M. Farajtabar, A. Ahmed, A. J. Smola, L. Song, Dirichlet-hawkes processes with applications to clustering continuous-time document streams, in: Proceedings of the 21th ACM SIGKDD International Conference on Knowledge Discovery and Data Mining, 2015, pp. 219–228.

[15] H. Xu, M. Farajtabar, H. Zha, Learning granger causality for hawkes processes, in: International Conference on Machine Learning, 2016, pp. 1717– 1726.

[16] U. Upadhyay, A. De, M. G. Rodriguez, Deep reinforcement learning of marked temporal point processes, in: Advances in Neural Information Processing Systems, 2018, pp. 3168–3178.

[17] S. Li, S. Xiao, S. Zhu, N. Du, Y. Xie, L. Song, Learning temporal point processes via reinforcement learning, in: Advances in neural information processing systems, 2018, pp. 10781–10791.

[18] G. Mohler, J. Carter, R. Raje, Improving social harm indices with a modulated hawkes process, International Journal of Forecasting 34 (3) (2018) 431–439.

[19] A. Reinhart, J. Greenhouse, Self-exciting point processes with spatial covariates: modelling the dynamics of crime, Journal of the Royal Statistical Society: Series C (Applied Statistics) 67 (5) (2018) 1305–1329.

[20] N. Altieri, R. L. Barter, J. Duncan, R. Dwivedi, K. Kumbier, X. Li, R. Netzorg, B. Park, C. Singh, Y. S. Tan, et al., Curating a covid-19 data repository and forecasting county-level death counts in the united states, arXiv preprint 2005.07882.

[21] A. C. Miller, N. J. Foti, J. A. Lewnard, N. P. Jewell, C. Guestrin, E. B. Fox, Mobility trends provide a leading indicator of changes in sars-cov-2 transmission, medRxiv.

[22] A. L. Lloyd, Destabilization of epidemic models with the inclusion of realistic distributions of infectious periods, Proceedings of the Royal Society of London. Series B: Biological Sciences 268 (1470) (2001) 985–993.

[23] T. Obadia, R. Haneef, P.-Y. Boëlle, The r0 package: a toolbox to estimate reproduction numbers for epidemic outbreaks, BMC medical informatics and decision making 12 (1) (2012) 147.

[24] B. J. Cowling, M. S. Lau, L.-M. Ho, S.-K. Chuang, T. Tsang, S.-H. Liu, P.-Y. Leung, S.-V. Lo, E. H. Lau, The effective reproduction number of pandemic influenza: prospective estimation, Epidemiology (Cambridge, Mass.) 21 (6) (2010) 842.

[25] J. Hellewell, S. Abbott, A. Gimma, N. I. Bosse, C. I. Jarvis, T. W. Russell, J. D. Munday, A. J. Kucharski, W. J. Edmunds, F. Sun, et al., Feasibility of controlling COVID-19 outbreaks by isolation of cases and contacts, The Lancet Global Health.

[26] A. L. Bertozzi, E. Franco, G. Mohler, M. B. Short, D. Sledge, The challenges of modeling and forecasting the spread of covid-19, Proceedings of the National Academy of Sciences 117 (29) (2020) 16732–16738. arXiv:https://www.pnas.org/content/117/29/16732.full.pdf, xdoi: 10.1073/pnas.2006520117. URL https://www.pnas.org/content/117/29/16732

[27] E. Lewis, G. Mohler, A nonparametric em algorithm for multiscale hawkes processes, Journal of Nonparametric Statistics 1 (1) (2011) 1–20.

[28] F. P. Schoenberg, Facilitated estimation of etas, Bulletin of the Seismological Society of America 103 (1) (2013) 601–605.

[29] J. Wallinga, P. Teunis, Different epidemic curves for severe acute respiratory syndrome reveal similar impacts of control measures, American Journal of epidemiology 160 (6) (2004) 509–516.

[30] S. Cauchemez, P.-Y. Boëlle, C. A. Donnelly, N. M. Ferguson, G. Thomas, G. M. Leung, A. J. Hedley, R. M. Anderson, A.-J. Valleron, Real-time estimates in early detection of sars, Emerging infectious diseases 12 (1) (2006) 110.

[31] The new york times, coronavirus in the u.s.: Latest map and case count (Mar 2020). URL https://www.nytimes.com/interactive/2020/us/coronavirus-us-cases.html

[32] E. Dong, H. Du, L. Gardner, An interactive web-based dashboard to track covid-19 in real time, The Lancet Infectious Diseases.

[33] Google, Covid-19 community mobility report (2020). URL https://www.google.com/covid19/mobility/

[34] K. Järvelin, J. Kekäläinen, Cumulated gain-based evaluation of ir techniques, ACM Transactions on Information Systems (TOIS) 20 (4) (2002) 422–446.

[35] D. Donoho, 50 years of data science, Journal of Computational and Graphical Statistics 26 (4) (2017) 745–766.

